# Cognitive Outcomes After Stenting vs. Endarterectomy: A Systematic Review with Meta-Analysis

**DOI:** 10.64898/2026.05.04.26351899

**Authors:** William J.P Ertl, Joshua Ward, Zachary A. Twomey, Francisco Call-Orellana, Uttam Verma, Shyian Jen, Hakeem Shakir

## Abstract

**Background:** Carotid artery stenosis may contribute to cognitive impairment through chronic hypoperfusion and subclinical ischemic injury. Although carotid endarterectomy (CEA) and carotid artery stenting (CAS) reduce stroke risk, their cognitive effects remain unclear. We conducted a systematic review and meta-analysis to evaluate cognitive outcomes after these interventions.

**Methods:** Following PRISMA guidelines, we searched PubMed, Embase, Web of Science, and the Cochrane Library from January 2000 to November 2025. Eligible studies reported cognitive outcomes after CEA or CAS, either alone or in direct comparison. Random-effects meta-analyses were performed for Mini-Mental State Examination (MMSE) and Montreal Cognitive Assessment (MoCA) outcomes. Single-arm studies were analyzed using standardized mean change, and head-to-head studies using mean difference. Outcomes were stratified by intervention type and follow-up interval (0–6 months and >6 months). Domain-specific cognitive outcomes were summarized qualitatively. Risk of bias was assessed using RoB-2, ROBINS-I, and the Newcastle-Ottawa Scale.

**Results:** Sixty-eight studies including 4,659 patients met inclusion criteria; 27 contributed to meta-analysis and 41 to qualitative synthesis. MMSE showed no significant early change after either intervention, while CAS showed significant improvement at >6 months. MoCA improved significantly after both CEA and CAS at early and late follow-up, although heterogeneity was high. Head-to-head analyses found no significant difference between CEA and CAS for MMSE or MoCA, but these comparisons were limited by small sample sizes. Domain-specific outcomes were mostly stable, with improvements most often reported in memory, attention, executive function, and processing speed.

**Conclusions:** Carotid revascularization may be associated with improved cognitive outcomes, particularly on MoCA, but results are heterogeneous and largely observational. Comparative evidence does not show a clear cognitive advantage of CEA or CAS. Future studies should use standardized cognitive testing and adequately powered direct comparisons.

## Introduction

Dementia is a clinical diagnosis characterized by progressive decline in cognitive function, often associated with neurodegeneration and cerebrovascular pathologies. It encompasses several major subtypes, which differ in predominant cognitive profiles, associated neuropsychiatric features, and underlying neural changes. Management strategies differ across these subtypes; however, vascular dementia—a major contributor to late-life cognitive impairment—is uniquely influenced by underlying cerebrovascular pathology. Many modifiable risk factors (e.g. chronic hypertension, diabetes mellitus, autoimmune disease) are shown to disrupt vascular function and cause underlying hemodynamic sources of impairment (e.g. carotid artery stenosis), predisposing patients to cognitive dysfunction.

Of the underlying hemodynamic sources of impairment, carotid artery stenosis is increasingly recognized as a major risk factor to vascular dementia through several mechanisms that promote improper cerebral perfusion, leading to gradual or sudden cognitive deficits.^1,2,3,4,5,6^ When cognitive decline results from ischemic injury, aspects of the impairment may be modifiable and display stabilization or partial recovery after correction of the underlying impairment.^2,3,7^

Treatment of carotid artery stenosis aims to restore proper tissue perfusion, depending on the severity and symptomatic status of the patient. Mild to moderate asymptomatic cases are managed with anti-platelet therapy and statins, however those with severe stenosis (>70%) or have focal neurologic symptoms (ex. stroke, TIA) require invasive interventions such as carotid endarterectomy (CEA) or carotid artery stenting (CAS). Both procedures are the most common revascularization techniques used for the prevention and treatment of stroke in patients with carotid stenosis ^7,8,9^, reestablishing blood flow to the brain parenchyma and restoring perfusion pressure.^3,4^

Prior studies examining cognitive outcomes after revascularization have produced inconsistent findings. Some authors report postoperative cognitive improvement, particularly in individuals with baseline hypoperfusion or impaired cerebrovascular reactivity, whereas others report minimal change or transient early postoperative decline.^10,11,12,13^ The wide array of cognitive outcome assessments have the additional effect of preventing significant comparison between individual studies adopting different standards of cognitive assessment, limiting conclusion of significant results.^14,15,16^

The goal of this systematic review is to evaluate changes in global cognitive function before and after carotid stenting or endarterectomy. By reporting results on global cognition as a clinically meaningful outcome and summarizing domain-specific testing outcomes all on an intervention-based assessment, this analysis integrates diverse assessments of cognition to characterize magnitude and direction of change within each procedure and summarize comparative evidence between stenting and endarterectomy.

## Methods

### Search Strategy

This systematic review followed PRISMA guidelines. PubMed, Embase, Web of Science, and Cochrane Library were searched from January 2000 to November 2025 using the search string with Boolean terms: (“carotid stenosis” OR “carotid artery disease”) AND ((“carotid stenting” OR “CAS”) OR (“carotid endarterectomy” OR “CEA”)) AND (“cognition” OR “dementia” OR “neuropsychological tests”).

### Study Selection and Data Extraction

Studies were included if they met the following criteria: adults ≥18 years with carotid artery stenosis undergoing CEA, CAS or comparing both, reporting at least one validated cognitive outcome, including global screening tests (Mini-Mental State Examination [MMSE] or Montreal Cognitive Assessment [MoCA]) or domain-specific assessments (e.g., memory, language, cognitive speed). We included randomized controlled trials, prospective cohorts, retrospective cohorts, and observational studies. Studies were excluded if they were case reports, review articles, editorials, abstracts, animal studies, ongoing or incomplete clinical trials, non-English publications, or lacked reporting of cognitive outcomes.

Two independent reviewers (W.E. and Z.T.) screened titles and abstracts using rayyan.ai. Conflicts were resolved by consensus of two additional reviewers (J.W. and F.C.O.). Full-text review was performed for all studies meeting preliminary criteria. Reasons for exclusion were recorded and documented in the PRISMA flow diagram (Figure 1).

**Figure 1.**
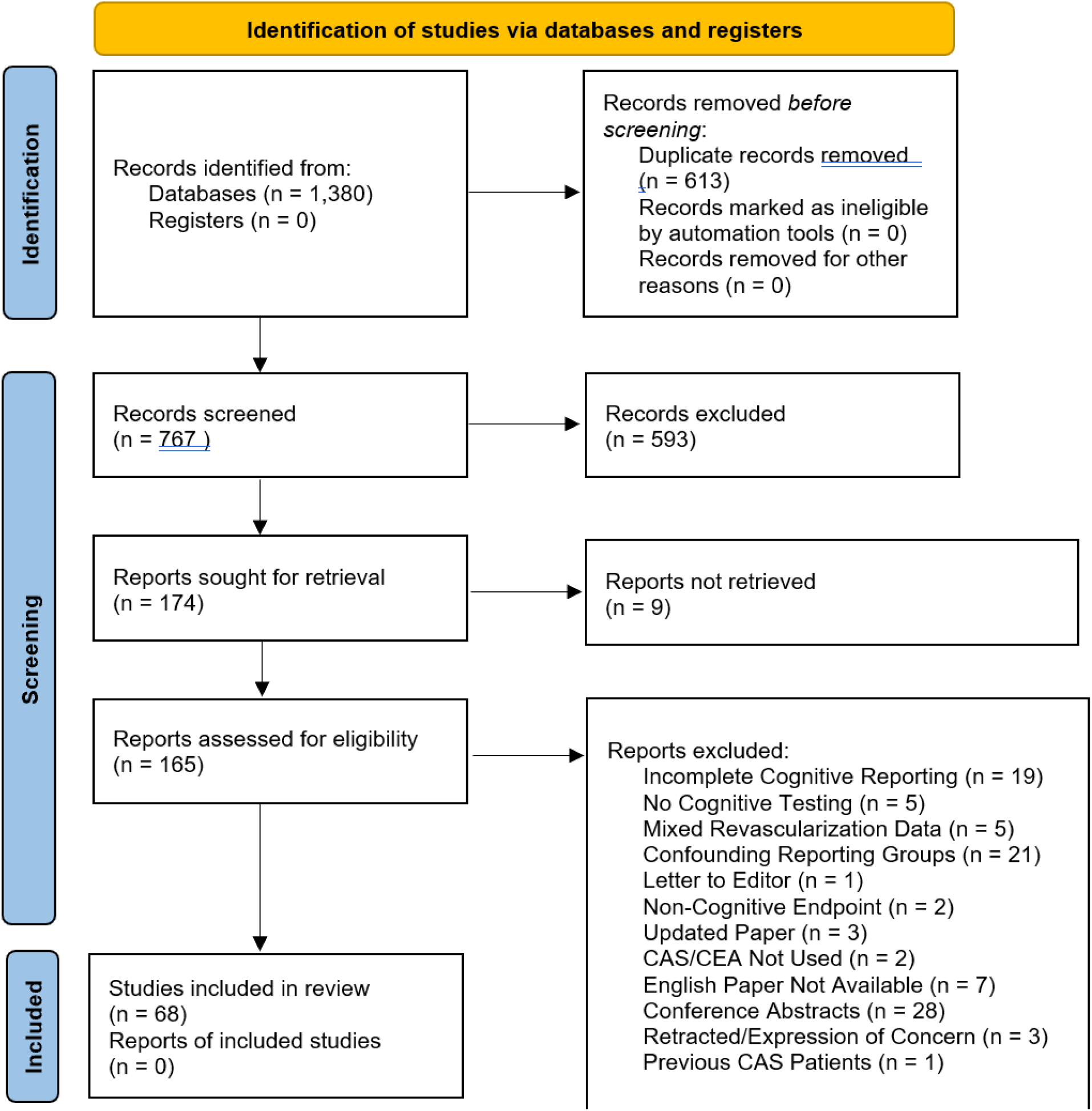
PRISMA diagram.

**Figure 2.**
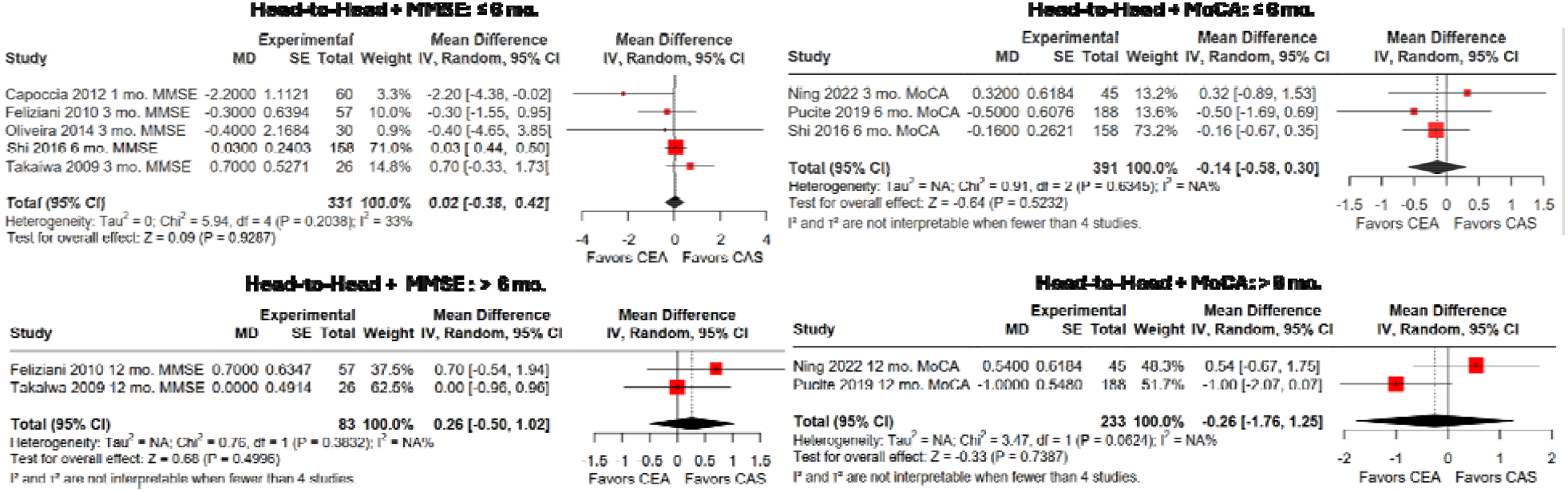
Head-to-head meta-analysis of cognitive outcomes following carotid revascularization. Forest plots show pooled differences in change from baseline scores (CAS − CEA) for MMSE and MoCA at early (0-6 months) and late (>6 months) follow-up. Positive values favor CAS. No statistically significant differences in cognitive change were observed between revascularization strategies

### Data Synthesis and Risk of Bias Assessment

Quantitative synthesis was performed for global cognitive outcomes assessed by MMSE and MoCA, as these are clinically relevant, widely used and commonly reported screening tools of global cognition. Domain-specific cognitive outcomes suffer from heterogeneity in reporting, instruments used, and scoring methodologies, therefore these were synthesized qualitatively based on study design and categorized as improvement, no change, or decline.

Risk of bias was evaluated by two reviewers (W.E. and Z.T.) using RoB-2 for randomized trials, ROBINS-I for non-randomized comparative studies, and the Newcastle–Ottawa Scale (NOS) for single-arm studies.

### Statistical Analysis

Two complementary analytic approaches were employed. Single-arm meta-analyses were conducted separately for CEA and CAS to estimate within-group change in global cognition. Head-to-head meta-analyses were performed using studies that directly compared CEA and CAS, estimating between-group differences in cognitive change. Analyses were based on change from baseline for MMSE and MoCA scores at two time points: early (≤ 6 months) and late (> 6 months). To avoid double-counting participants, only one representative time point per study per interval was included. When multiple assessments were reported within an interval, outcomes closest to 3 months and 12 months were selected for early and late follow-up, respectively.

Cognitive change was defined as the mean change between post- and pre-procedure scores. Mean change was used as the effect size metric for single-arm analyses (CAS or CEA), whereas mean difference was used for head-to-head studies (CAS & CEA). When the standard deviation of the change score was not reported, it was estimated from pre- and post-procedure standard deviations assuming a within-subject correlation coefficient of 0.5, consistent with established practice in longitudinal meta-analyses. Study arms with non-informative (zero) variance were excluded from quantitative synthesis. All meta-analyses were conducted using random-effects models fitted with restricted maximum likelihood (REML) estimation to account for between-study heterogeneity. Pooled estimates are reported with 95% confidence intervals. Statistical heterogeneity was assessed using τ^2^ and I^2^ statistics.

When studies reported outcomes for clinically distinct subgroups (e.g., symptomatic versus asymptomatic patients), each subgroup was treated as an independent cohort to preserve clinically meaningful distinctions. Sensitivity analyses were performed to assess the robustness of the findings, including restriction to a single follow-up time point per interval and exclusion of cohorts with non-informative variance. Head-to-head analyses based on a limited number of contributing studies were interpreted cautiously and considered exploratory.

Small-study effects were assessed with funnel plots (effect estimate versus standard error) using two tests: a linear regression test of funnel plot asymmetry (Egger-type) and a rank correlation test (Begg–Mazumdar type). Plots were constructed using study effect estimates against their standard errors (precision), consistent with the random-effects meta-analytic models. Trim-and-fill was used as a sensitivity imputation; it is not proof of missing studies. Funnel plots and asymmetry tests were performed separately for: (i) direct comparative (double-arm) CAS-versus-CEA analyses, (ii) single-arm CAS cohorts, and (iii) single-arm CEA cohorts. This approach was chosen to align bias assessment with study design and intervention context as instrument-specific asymmetry testing was not the primary bias objective.

Following meta-analysis, single-arm subgroups meeting prespecified criteria (≥ 5 effect sizes and I^2^ > 50%; n = 11 analyses) were subjected to meta-regression to evaluate the influence of commonly reported covariates, specifically symptomatic status and baseline cognitive function. Meta-regression models were fitted using random-effects with REML estimation and regression coefficients (β) to quantify associations between covariates and pooled effect sizes. Because symptomatic status definitions and inclusion criteria varied, analyses were restricted to studies that clearly defined individual covariants. Model fit was summarized using R^2^, and influence and residual diagnostics were evaluated using Cook’s distance and the Shapiro–Wilk test.

Meta-analyses and regression were performed in R (version 4.5.2) using the *metafor* package^17^. All statistical tests were two-sided, with p < 0.05 considered statistically significant.

## Results

### Study Selection, Overview, and Study Population

Out of the 1,380 studies assessed, 68 met our inclusion criteria (Figure 1). 27 studies were included in the meta-analyses while the remaining 41 underwent qualitative assessment only. There were 4,659 enrolled patients, of whom 4,633 underwent formal cognitive assessment (Table 1). Median reported age was 68.8 years (range 57–77), and 73.2% of participants were male. Eighteen studies evaluated patients undergoing CEA and 29 evaluated CAS, with the remaining studies including mixed procedural cohorts. Most studies enrolled symptomatic and asymptomatic populations; among explicitly defined studies, 71.1% of patients were asymptomatic. Median follow-up duration across all studies was 6 months (range 0.5–216 months). Cognitive assessment strategies varied considerably; MMSE and MoCA used in 31 and 16 studies while domain-specific neuropsychological testing was employed in the majority of studies (59/68). Study characteristics are summarized in Table 1. Individual study demographics are included in Supplementary Table 1.

**Table 1.** Study demographics and baseline characteristics. Summary of study-level demographic and clinical characteristics for all included studies stratified by single arm CEA, single arm CAS, and studies including CEA and CAS.

| Table 1. Study Demographics |  |  |  |  |
| --- | --- | --- | --- | --- |
| Characteristic | Total | CEA | CAS | CEA/CAS |
| Number of studies (k) | 68 | 18 | 29 | 21 |
| Total patients (N) | 4659 (enrolled); 4633 (cognitive cohort) | 1193 (enrolled); 1190 (cognitive cohort) | 1657 (enrolled); 1634 (cognitive cohort) | 1809 (enrolled); 1809 (cognitive cohort) |
| Age, years (median [IQR] or range) | 68.8 [67–71.3]; range 57–77 | 68.8 [67.8–72.7]; range 57–77 | 68.4 [66–71]; range 60–76 | 68.8 [67.6–70.8]; range 64.9–75.6 |
| % male | 73.2% (range 52.3–100) | 76.1% (range 56.7–92.3) | 73.6% (range 52.3–100) | 69.8% (range 56–87.2) |
| Symptomatic status | Asymptomatic (23); Mixed (35); Not reported (3); Symptomatic (7) | Asymptomatic (5); Mixed (11); Not reported (1); Symptomatic (1) | Asymptomatic (10); Mixed (13); Not reported (1); Symptomatic (5) | Asymptomatic (8); Mixed (11); Not reported (1); Symptomatic (1) |
| % asymptomatic | 71.1% (counts in 49 studies) | 66.7% (counts in 10 studies) | 71.5% (counts in 22 studies) | 72.7% (counts in 17 studies) |
| % symptomatic | 28.9% (counts in 49 studies) | 33.3% (counts in 10 studies) | 28.5% (counts in 22 studies) | 27.3% (counts in 17 studies) |
| Follow-up duration | 6 [3–12] months; range 0.5–216 | 4.5 [1.2–12] months; range 0.5–216 | 6 [3–6] months; range 2–27.6 | 4.5 [3–12] months; range 1–12 |
| Cognitive assessment used | MMSE: 31/68 studies; MoCA: 16/68 studies; Domain-specific tests: 59/68 studies | MMSE: 8/18 studies; MoCA: 4/18 studies; Domain-specific tests: 14/18 studies | MMSE: 14/29 studies; MoCA: 8/29 studies; Domain-specific tests: 29/29 studies | MMSE: 9/21 studies; MoCA: 4/21 studies; Domain-specific tests: 16/21 studies |

#### Single-arm meta-analyses

Studies evaluating MoCA exhibited higher mean change values than MMSE at early and late follow-up. Results from single-arm and head-to-head analyses are explained in the following subsections and summarized in Table 2, with corresponding forest plots shown in Figures 3-4. Sensitivity analyses demonstrated the observed cognitive effects were robust to correlation assumptions. Pre-post correlation variation (0.3, 0.5, and 0.8) did not materially alter effect estimates, confidence intervals or statistical significance.

**Table 2.** Random-effects meta-analysis of MMSE and MoCA outcomes after CEA and CAS, stratified by follow-up interval. Single-arm analyses summarize within-group change from baseline, and head-to-head analyses compare CAS versus CEA directly. Values shown are pooled estimates with 95% confidence intervals; k indicates number of included study arms or comparisons and N the total sample size.

| Analysis | k | N | Estimate | 95% CI | p value | I<sup>2</sup> (%) |
| --- | --- | --- | --- | --- | --- | --- |
| CEA MMSE (0-6 mo) | 11 | 403 | 0.51 | [-0.14, 1.16] | 0.1258 | 0.74 |
| CEA MMSE (>6 mo) | 5 | 197 | 0.59 | [-0.06, 1.23] | 0.0731 | 0.71 |
| CEA MoCA (0-6 mo) | 5 | 401 | 0.81 | [0.51, 1.11] | 1.6e-07 | 0.59 |
| CEA MoCA (>6 mo) | 4 | 324 | 1.33 | [0.53, 2.13] | 0.0011 | 0.92 |
| CAS MMSE (0-6 mo) | 13 | 586 | 0.38 | [-0.28, 1.04] | 0.2569 | 0.85 |
| CAS MMSE (>6 mo) | 5 | 114 | 1.12 | [0.50, 1.73] | 4e-04 | 0.53 |
| CAS MoCA (0-6 mo) | 8 | 430 | 2.59 | [0.54, 4.63] | 0.0133 | 0.97 |
| CAS MoCA (>6 mo) | 4 | 138 | 2.93 | [1.56, 4.30] | 2.8e-05 | 0.90 |
| Head-to-head MMSE (0-6 mo) | 5 | 331 | 0.02 | [-0.38, 0.42] | 0.9287 | 0.33 |
| Head-to-head MMSE (>6 mo) | 2 | 83 | 0.26 | [-0.50, 1.02] | 0.4996 | N/A |
| Head-to-head MoCA (0-6 mo) | 3 | 391 | -0.14 | [-0.58, 0.30] | 0.5232 | N/A |
| Head-to-head MoCA (>6 mo) | 2 | 233 | -0.26 | [-1.76, 1.25] | 0.7387 | N/A |

**Figure 3.**
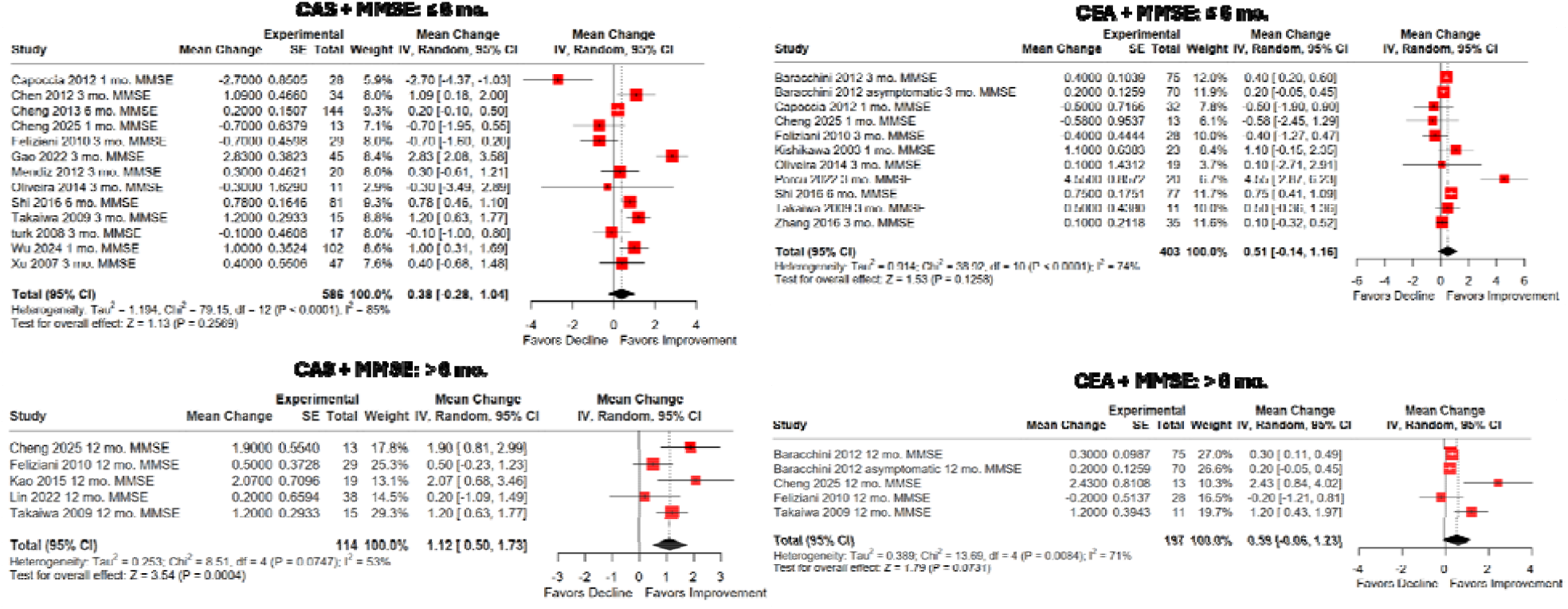
Single arm meta-analysis outcomes of MMSE. Forest plots illustrate pooled within-group changes in MMSE scores from baseline following CEA or CAS, stratified by early (0-6 months) and late (>6 months) follow-up. Random-effects models were applied. Positive changes represent improvement relative to baseline.

**Figure 4.**
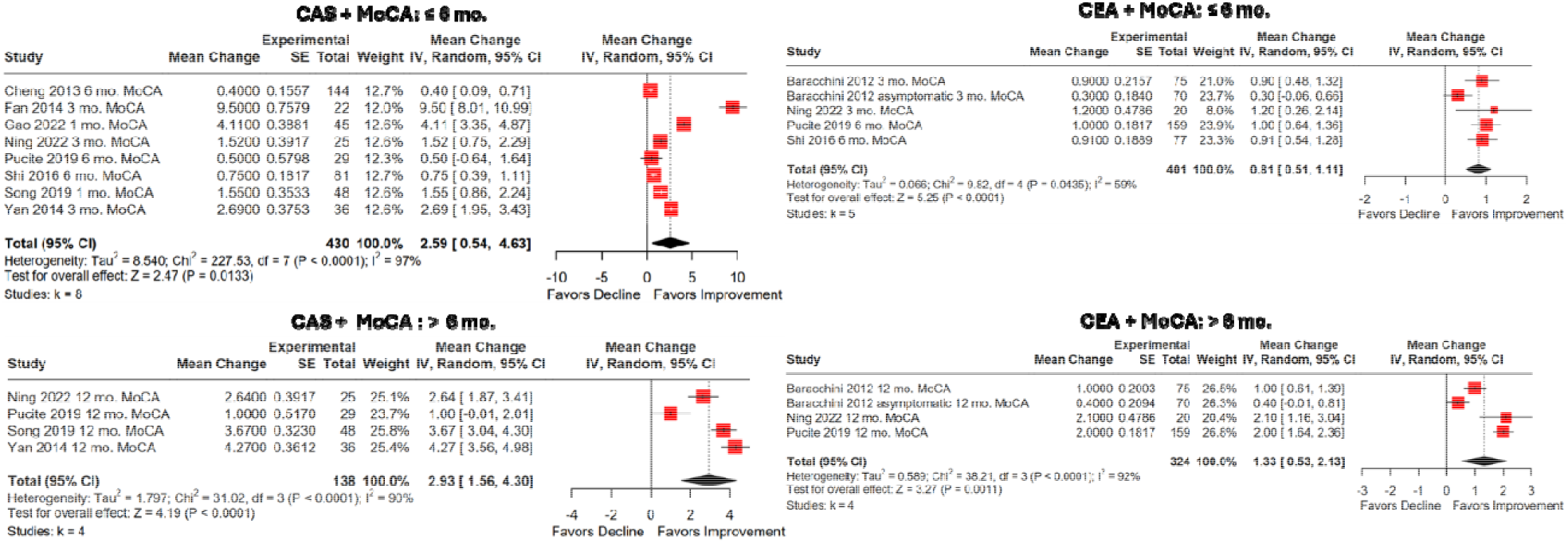
Single arm meta-analysis of MoCA. Forest plots show pooled within-group changes of MoCA scores from baseline following CEA or CAS, stratified by early (0-6 months) and late (>6 months) follow-up. Random-effects models were applied. Positive changes represent improvement relative to baseline.

### Carotid Endarterectomy and Global Cognition

CEA was associated with positive mean change in global cognitive scores. On the MMSE, pooled change scores were 0.51 at ≤ 6 months (95% CI −0.14 to 1.16) and 0.59 at > 6 months (95% CI −0.06 to 1.23). MoCA scores showed larger mean change, with pooled increases of 0.81 points at ≤6 months (95% CI 0.51 to 1.11) and 1.33 at >6 months (95% CI 0.53 to 2.13). Detailed study characteristics, heterogeneity measures, and sample sizes are reported in Table 2.

### Carotid Artery Stenting and Global Cognition

For CAS, MMSE scores demonstrated a mean change of 0.38 at ≤6 months (95% CI −0.28 to 1.04) and 1.12 points at >6 months (95% CI 0.50 to 1.73). MoCA scores showed larger mean change at both time points, with pooled increases of 2.59 at ≤ 6 months (95% CI 0.54 to 4.63) and 2.93 at > 6 months (95% CI 1.56 to 4.30). Corresponding heterogeneity estimates and study counts are summarized in Table 2.

#### Head-to-head meta-analysis

Direct comparisons between CAS and CEA were limited due to a low number of studies comparing both interventions. Pooled effect differences were 0.02 (95% CI -0.38, 0.42) and - 0.26 (95% CI -1.76, 1.25) for MMSE at ≤ 6 months and > 6 months, respectively. Pooled effect differences for MoCA were -0.14 (95% CI -0.58, 0.30) and 0.26 (95% CI -0.50, 1.02) for ≤ 6 months and > 6 months, respectively. No statistically significant findings were found from any of the analyses (all values of p > 0.5). Although head-to-head studies at ≤ 6 months using MMSE yielded low heterogeneity (I^2^ = 33%), small study sizes in all other subgroups limited heterogeneity assessment (k ≤ 4), so heterogeneity analysis was not performed on these subgroups. Head-to-head forest plots are shown in Figure 2.

#### Publication Bias Assessment

Overall, formal funnel-asymmetry tests were non-significant across both double-arm and single-arm groupings, providing no strong statistical signal of pervasive small-study asymmetry in this dataset. All tests, excluding CAS ≤ 6 months and CEA ≤ 6 months, contained a low number of included studies (*k*), limiting analysis to exploratory understanding. Funnel plots are shown in Supplemental Figure 2

### Head-to-head Analysis

At ≤ 6 months (*k* = 8; *N* = 722), neither the Egger-type test (*p* = 0.454) nor the Begg-type test (*p* = 0.548) suggested significant funnel asymmetry. Trim-and-fill imputed 2 studies, with a small shift in pooled contrast from -0.054 to 0.018. Between-study variance was low (τ^2^ = 0.000). At > 6 months (*k* = 4; *N* = 316), neither test was significant (Egger-type *p* = 0.470; Begg-type *p* = 0.333), and trim-and-fill imputed no studies; the pooled contrast was unchanged (0.016). τ^2^ was 0.249.

### Endarterectomy

At ≤ 6 months (*k* = 16; *N* = 804), asymmetry tests were non-significant (Egger-type *p* = 0.430; Begg-type *p* = 0.825), with no trim-and-fill imputations; pooled change remained 0.599. τ^2^ was 0.266. At > 6 months (*k* = 9; *N* = 521), tests were non-significant (Egger-type *p* = 0.198; Begg-type *p* = 0.358). Trim-and-fill imputed 1 study, shifting pooled change from 0.965 to 0.864. τ^2^ was 0.622.

### Stenting

At ≤ 6 months (*k* = 21; *N* = 1,016), asymmetry tests were non-significant (Egger-type *p* = 0.211; Begg-type *p* = 0.976), but trim-and-fill imputed 6 studies, with a larger shift in pooled change from 1.176 to 1.872. τ^2^ was high (4.878), indicating substantial heterogeneity. At > 6 months (*k* = 9; *N* = 252), tests were non-significant (Egger-type *p* = 0.510; Begg-type *p* = 0.919), trim-and-fill imputed no studies, and pooled change was unchanged (1.972). τ^2^ was 1.790.

#### Regression Analysis

Meta-regression analyses were conducted on single-arm subgroups meeting prespecified criteria. Baseline cognitive function was a significant negative moderator of cognitive change in three subgroups: CEA-MMSE ≤6 months (β = −0.382, p < .001, R^2^ = 100%), CEA-MMSE ≤6 months asymptomatic (β = −0.476, p < .001, R^2^ = 100%), and CAS-MoCA ≤6 months (β = −0.591, p = .011, R^2^ = 67.39%). Symptomatic status was significantly moderated cognitive change in CEA-MoCA ≤ 6 months (symptomatic vs asymptomatic: β = 0.627, p = .023, R^2^ = 100%), while papers reporting mixed symptom status were also significant in CEA-MoCA >6 months (mixed vs asymptomatic: β = 1.484, p = .033, R^2^ = 100%). CAS-MMSE ≤ 6 months mixed showed a significant positive baseline effect (β = 0.396, p = .010, R^2^ = 100%). No moderators were statistically significant in the remaining CAS-MMSE subgroups (R^2^ ≤ 6%) or CAS-MoCA ≤6 months mixed (p = .580).

Influence diagnostics were conducted for the single-arm effect sizes. Cook’s distance was used to identify studies with undue influence on the regression coefficients; the cut-off for influence was the 50% quantile of a chi-square distribution with 4 degrees of freedom (3.357). 1 effect size(s) exceeded this cut-off (Fan 2014, 3 mo.). Sensitivity analysis excluding this effect size yielded similar coefficient estimates and inference. Although the Shapiro–Wilk test suggested deviation from normality (W = 0.93, p = .015), visual inspection of residual plots did not reveal substantial departures from linearity or homoscedasticity, and the model was judged to provide an adequate approximation to the data.

#### Domain Specific Cognitive Outcomes

Cognitive outcomes in domain-specific testing most frequently demonstrated improvement or stability, with decline scarcely reported. By study design, improvements were most numerically reported in memory, global cognition, attention, and processing speed in studies only evaluating CAS; global cognition and memory for CEA only studies; and in language, memory, attention, and executive function for head-to-head studies. Reported declines were uncommon across all studies and typically limited to isolated domains, individual tests, or study-design based. The qualitative synthesis can be visualized in Figure 5.

**Figure 5.**
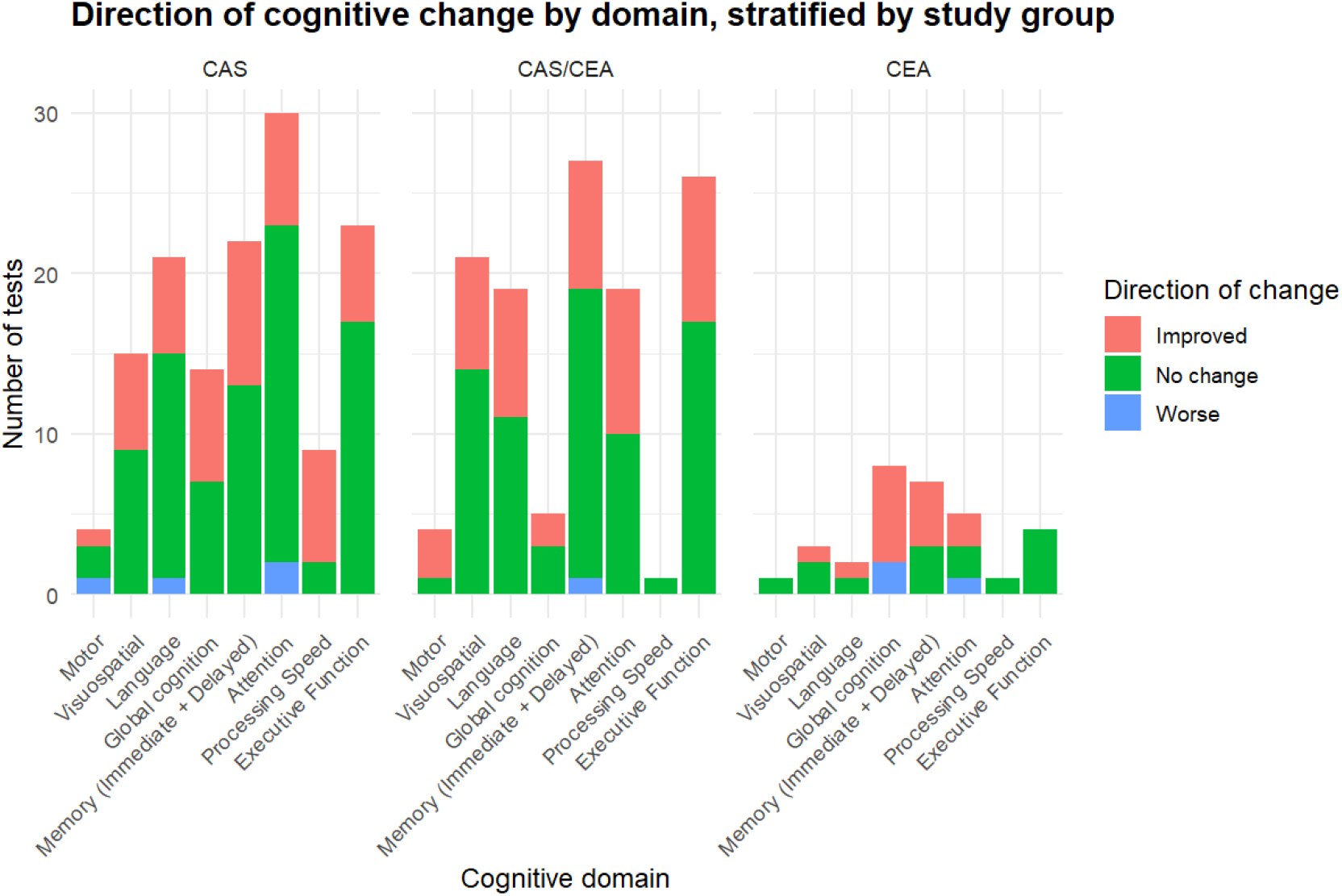
Direction of cognitive change by domain, stratified by study group. Stacked bar plots show the number of cognitive tests demonstrating improvement, no change, or decline across cognitive domains following CAS, CEA and studies including both interventions (CAS/CEA).

#### Risk of Bias

Of the studies assessed, most were assessed using the Newcastle–Ottawa Scale (n = 45, 67%) with the rest falling into the ROBINS-I (n = 20, 29%) or RoB-2 (n = 3, 4%) based on study design. Most studies were assessed to be either of moderate/some concerns (67.6%) to high/serious (27.9%) risk of bias, with the rest being low risk. Risk-of-bias summaries are presented in Table 3, with study-level assessments provided in Supplementary Table 2.

**Table 3.** Risk of bias summary by study design. Overall risk of bias across included studies, stratified by study design and risk-of-bias assessment tool. Risk categories were harmonized descriptively across tools and ordered from lowest to highest risk to facilitate visual comparison; categories are not intended to imply direct equivalence between instruments. Tool specific descriptions are described in the text. Values are reported as the number of studies with corresponding percentages within each study design.

| Risk of bias summary by study design |  |  |  |  |  |  |  |
| --- | --- | --- | --- | --- | --- | --- | --- |
| Study design | Risk-of-bias tool | Studies, n | Low | Moderate | High | Serious | Some concerns |
| Cohort studies | NOS | 45 | 2 (4.4%) | 35 (77.8%) | 8 (17.8%) | 0 (0.0%) | 0 (0.0%) |
| Nonrandomized comparative studies | ROBINS-I | 20 | 1 (5.0%) | 8 (40.0%) | 0 (0.0%) | 11 (55.0%) | 0 (0.0%) |
| Randomized trials | RoB-2 | 3 | 0 (0.0%) | 0 (0.0%) | 0 (0.0%) | 0 (0.0%) | 3 (100.0%) |

## Discussion

This systematic review and meta-analysis synthesized changes in global cognitive outcomes following CEA and CAS across studies pooling patients with carotid artery stenosis. Single-arm analysis of all interventions and scales yielded cognitive improvement except in CAS + MMSE ≤ 6 months. However, heterogeneity remained moderate to high in all comparisons (I^2^ > 50%) due to unaccounted confounding factors.

In contrast, head-to-head comparisons of both MMSE and MoCA demonstrated no significant between-procedure difference in change scores CEA at either time point, with confidence intervals reflecting limited comparative evidence.

Of note, CAS MoCA pooled changes (PC = 2.59 at ≤ 6 months, 2.93 at > 6 months) were dramatically larger than CEA MoCA pooled changes (PC = 0.81 at ≤ 6 months, 1.33 at > 6 months). These differences are most likely the result of distinct sets of studies with differing designs and patient mixes, reflecting the bodies of evidence summarized for each revascularization strategy—including study number, heterogeneity, and reporting—rather than a superior cognitive recovery relative to endarterectomy independent of these contextual factors.

Overall, clinically significant changes based on minimum clinically significant difference (MCID) estimates were most observed in studies evaluating cognition using MoCA. Greater than or equal to 1 point of change is considered a good indicator in MoCA for a MCID^33^, present in every single-arm analysis except CEA ≤ 6 months (Pooled Change (PC) = 0.81). A robust indicator for MCID in MMSE is generally considered ≥ 2 points of change, although a 1-point change can sometimes reflect a minimal clinically important difference depending on daily fluctuation and baseline severity of cognitive impairment/dementia^34,35^. While CAS > 6 mo did achieve a MCID (PC = 1.12) on the MMSE, moderate heterogeneity (I^2^ = 53%) in addition to minimal studies included (k = 5) should caution meaningful interpretation.

Our qualitative synthesis of domain-specific outcomes reflected overall study design variability across studies. The wide array of testing batteries, scoring approaches, and follow-up timing limits the interpretation and clinical significance of the quantitative analysis. However, some of the domains most frequently reported as improved (memory, attention, and executive function) align with cognitive profiles described in vascular cognitive impairment and with pathophysiological models of hypoperfusion and impaired cerebrovascular reserve.^18,19,20^

Across both comparative and within-procedure evidence families, formal funnel asymmetry tests did not detect statistically significant asymmetry: however, interpretation should not rely on *p*-values alone. Several analyses had small *k*, and heterogeneity ranged from low to substantial depending on subgroup. High heterogeneity in CAS single-arm analyses (especially ≤6 months) complicates inference and may contribute to funnel shape independent of selective reporting. Trim-and-fill sensitivity was mostly stable in some strata (e.g., no or minimal imputation effects in several analyses), but notable shifts occurred in selected groups, particularly CAS single-arm ≤6 months, emphasizing cautious interpretation of single-arm pooled improvements.

Meta-regression analyses performed on single-arm analyses revealed symptomatic status as a significant positive moderator in CEA-MoCA subgroups. This patterns suggest greater cognitive gains after CEA among patients with lower baseline scores and symptomatic presentations with ample study size and moderate heterogeneity (k ≥ 5 studies with I^2^ > 50%). However, this result should be interpreted cautiously; MoCA has higher sensitivity to small changes in cognitive function when compared with MMSE, and residual heterogeneity remains substantial in CAS-MMSE analyses. Additionally, several models reported R^2^ = 100%: this is atypical and reflects overfitting as opposed to model perfection from limited subgroup studies included.

Several biological mechanisms may explain why cognitive changes after revascularization are heterogeneous. Chronic carotid stenosis can impair cognition through impaired cerebral perfusion and reduced cerebrovascular reserve, even in the absence of overt infarction and potentially affecting the domains suggested by Mendiz et al and Lal et al.^18,19,20^. Despite the benefits of revascularization, additional periprocedural factors (such as microembolization, anesthesia side effect, and postoperative variability) may contribute to transient early decline in some individuals. ^19,21,22,23^ Additionally, cognition as well as dementia have added systemic and regional factors that can contribute towards decline. Although these are out of scope of our study, they may have significant contributions toward clinical presentation/outcome and must be taken into management consideration on a case-by-case basis.

Differences between MMSE and MoCA outcomes, in our study and across the literature, likely reflect measurement characteristics. MMSE was designed originally as a brief assessment of global cognition and may be less sensitive to mild cognitive deficits, while MoCA includes more executive and attentional components and has a higher ability to discriminate normal cognition from mild cognitive impairment (MCI) and early onset dementia than previous tools. In the context of carotid stenosis, these properties may influence the likelihood of detecting small changes after intervention; however, greater sensitivity may also increase susceptibility to practice effects and to identify false-positives depending on the cutoff used and population characteristics.^24,25,26^

Direct comparisons of cognitive outcomes after CAS or CEA have been limited. Several prospective and observational studies have reported postoperative cognitive improvement following CEA or CAS, particularly among patients with baseline hypoperfusion or impaired cerebrovascular reactivity.^27,28,29^ However, cognitive effects in the peri- and postoperative period have been highly variable and appear to depend on study characteristics.^14,16,30,31^ Prior comparative literature has shown comparable long-term neurological outcomes between the two procedures, with higher-level studies focusing primarily on peri-procedural risk profiles rather than longitudinal cognitive endpoints.^8,10^ In this context, our head-to-head meta-analysis extends this by detailing available comparative cognitive data and showing no statistically significant cognitive advantage of one procedure over the other. However, limited studies comparing both interventions and wide confidence intervals limited precision, urging the need of adequately powered comparative studies with standardized interventions, tests and outcomes.

Recent stroke-outcomes trials in asymptomatic carotid stenosis, including CREST-2, underline how decisions are increasingly anchored on intensive medical therapy and selective revascularization in defined populations. Stroke-related outcomes from randomized trials and routine reporting systems do not substitute for cognitive endpoints measured with standardized neuropsychological tools. Therefore, comparisons based on stroke cannot settle whether, how much, or for whom cognition changes after carotid revascularization.^32^ The incorporation of standard cognitive endpoints into vascular outcomes can further clarify benefits in silent ischemia, impaired cerebrovascular reserve and mixed vascular neurodegenerative pathology.

### Limitations

Available literature on cognitive outcomes after carotid revascularization was highly heterogeneous with substantial variability in patient selection, design, periprocedural management, and cognitive assessment strategies between studies. Additionally, our analysis had no true control to compare outcomes due to limited reporting of controls in individual studies, making a comparison to best medical treatment nonsensical.

Inconsistent test selection, follow-up timing, and reporting of changes in scores meant meta analysis was restricted to the most commonly reported global screening instruments (MoCA and MMSE). This approach excludes other studies assessing global cognition with other individual tests (Mini-Cog, ACE-III, GPCOG, etc.), limiting our study selection. Additionally, domain specific outcomes were summarized qualitatively using direction of change categories without meaningful interpretation. This allowed for synthesis across instruments and should be interpreted as descriptive only.

The inclusion of both symptomatic and asymptomatic carotid stenosis populations represents an additional limitation. Baseline cognitive status, cerebrovascular reserve, and vulnerability to peri-procedural injury are likely to differ between these groups, yet these factors were inconsistently reported and adjusted for across studies.

Most included studies were observational, introducing potential confounding related to vascular comorbidities, educational attainment, baseline cognition, and treatment selection. Practice effects and regression to the mean may also have influenced observed improvements, particularly in studies lacking appropriate control groups or repeated baseline testing.

As noted earlier, follow-up timing was inconsistent across studies and cognitive trajectories after carotid revascularization may vary over time. To enable quantitative synthesis, follow-up intervals were divided into early and late time points. While this approach allows for cross-study comparison and prevents double counting, it sacrifices temporal granularity and can obscure short-term fluctuations.

This study was not registered to PROSPERO, an additional limitation to our study.

### Future Directions

More head-to-head studies comparing stenting to endarterectomy should be done before meaningful results can be drawn. Single and double-armed analysis should also incorporate standardized, longitudinal cognitive testing protocols with follow up intervals reflecting the immediate post-operative, short-term, and long-term cognitive effects of each intervention. Future studies should also outline true, standardized, non-surgical controls to prevent confounding in medical management. While some single arm studies included the use of best medical treatment, most commonly statins + P2Y12 inhibitor, many did not have a true control to compare outcomes. These changes can provide the adjustment needed for future studies investigating stenting, endarterectomy, and other techniques such as trans-carotid artery revascularization (TCAR) or investigational procedures like intravascular lithotripsy with angioplasty and stenting.

## Conclusion

Carotid revascularization was associated with measurable changes in global cognitive screening scores in pooled single-arm analyses, with MoCA generally appearing more responsive than MMSE across early and late follow-up. These within-procedure summaries should be interpreted cautiously given high between-study heterogeneity, predominantly observational designs, and the potential for practice effects and regression to the mean. In direct comparative analyses, we did not identify a statistically significant difference in cognitive change between carotid artery stenting and carotid endarterectomy; however, this evidence was sparse and imprecise, and non-significance should not be interpreted as procedural equivalence. Domain-level evidence remained heterogeneous and descriptive. Clarifying whether revascularization preserves or improves cognition will require prospective studies with standardized, longitudinal neuropsychological assessment, blinded testing where feasible, pre-specified cognitive endpoints adequately powered comparisons in well-defined symptomatic and asymptomatic populations.

## Supporting information

All Supplemental Tables

## Data Availability

All data contained within the manuscript is available upon request from the primary author

