## Supplementary material for "Cognitive Outcomes After Stenting vs. Endarterectomy: A Systematic Review with Meta-Analysis": All Supplemental Tables

**SUPPLEMENTAL TABLE 1.** Characteristics of studies included in the systematic review and meta-analysis of cognitive outcomes after CEA and CAS. N = total participants.

| Study | Design | Symptomatic | Arms | Scales | N | % male |
| --- | --- | --- | --- | --- | --- | --- |
| Aharon-  Peretz 2003 | non-RCT comparative | Asymptomatic | CEA | MMSE, MoCA, WAIS, TMT, Digit Span,  Corsi Blocks, RCF, RAVLT, Tower of London | 60 | 56.7 |
| Akkaya  2014 | non-RCT comparative | Asymptomatic | CAS | Rey auditory verbal, Digit span, TMT, Verbal fluency, Stroop, Rey complex figure | 100 | 57 |
| Baracchini  2012 | non-RCT comparative | Asymptomatic + Symptomatic, separated | CEA | MMSE, MoCA | 213 | 71.4 |
| Capoccia  2012 | non-RCT comparative | Asymptomatic | CEA,  CAS | MMSE | 60 | 63.3 |
| Carta 2015 | non-RCT comparative | Mixed | CEA | SF-12, Hamimlton. WAIS | 40 | 62.5 |
| Chen 2012 | Prospective | Asymptomatic | CAS | NIHSS, Barthel Index, ADAS, MMSE, TMT,  Verbal Fluency | 34 | 76 |
| Cheng 2013 | non-RCT comparative | Mixed | CAS | MMSE, MoCA, FOME, RVR, WAIS-DS,  ADL, NIHSS | 210 | 66.7 |
| Cheng 2025 | Prospective | Mixed | CEA | MMSE, MoCA | 13 | 92.3 |
| Er 2021 | Prospective | Mixed | CEA | Verbal Memory Process, WMS, Digit span,  Verbal fluency | 38 | 78.9 |
| Fan 2014 | non-RCT comparative | Symptomatic | CAS | NIHSS, MoCA | 40 | 87.5 |
| Feliziani  2010 | non-RCT comparative | Asymptomatic | CEA,  CAS | MMSE, ADL, IADL, Babcock SR, Rey-IR,  Rey-DR. CNT, TMT, COWA, CD | 46 | 67.4 |
| Gao 2022 | RCT | Symptomatic | CAS | MMSE, MoCA, Barthel Index | 90 | 58.9 |
| Grunwald  2010 | Prospective | Asymptomatic | CAS | CNT, Labyrinth, FST, CWT | 41 | 73.2 |
| Gupta 2020 | Prospective | Symptomatic | CAS | ACE | 28 | 89 |
| Huang 2013 | non-RCT comparative | Mixed | CAS | MMSE, NIHSS, Barthel Index, ADAS, Color  Trails, Verbal fluency | 59 | 72.9 |
| Huang 2018 | non-RCT comparative | Mixed | CAS | MMSE, MoCA, CVLT, Stroop, TMT-A,  Stroop | 30 | 70 |
| Ishihara  2013 | Prospective | N/A | CAS | WMS, WAIS-III | 39 | 94.9 |
| Kao 2015 | Prospective | Symptomatic | CAS | MMSE, NIHSS, Barthel Index, ADAS, Color  Trails, Verbal fluency | 19 | 78.9 |
| Kishikawa  2003 | non-RCT comparative | Mixed | CEA | MMSE, HDSR, Block-design, Verbal-memory | 40 | 90 |
| Kougias  2015 | non-RCT comparative | Asymptomatic | CEA,  CAS | WAIS-IV, Stroop, TMT, JLO, BVMT-R,  RAVLT, FAS, Category, Stroop, Grooved  Pegboard, Depression, Anxiety | 55 | N/A |
| Kramska  2022 | Prospective | Symptomatic | CEA | RBANS | 60 \|  Cognitive cohort: 57 | 72 |
| Kuliha  2014 | RCT | Mixed | CEA,  CAS | MMSE, Clock-drawing, Verbal fluency | 150 | 70 |
| Lal 2011 | non-RCT comparative | Asymptomatic | CEA,  CAS | TMT, BNT, Working Memory Test, COWA,  HVLT | 45 | 65.2 |
| Lehrner  2005 | Prospective | Mixed | CAS | AKT, TMT, WAIS, Verbal fluency, Stroop, c. I. | 20 | 70 |
| Lin 2016 | non-RCT comparative | Asymptomatic | CEA,  CAS | MMSE, Total immediate recall, Delayed recall, symbol digit modalities, M-TMT, Stroop,  Complex Figure | 40 | 80 |
| Lin 2022 | Non-RCT  Comparative | Asymptomatic | CAS | Delayed verbal memory, MMSE, TMT-A,  TMT-B, Stroop, DHI, Copy figure test, Recall figure test, Amnestic MCI, WM volumes, GM volumes, Hippocampi volumes, Scheltens rating scale | 69 \|  Cognitive cohort: 46 | 58 |
| Mendiz  2012 | Prospective | Asymptomatic | CAS | MMSE, ACE-R, BNT, Semantic fluency,  RAVLT, Rey complex figure, digit span, TMT, WCST, phonological flunecy, IFS, digit symbol, symbol search | 20 | 65 |
| Miyamatsu  2022 | Retrospective | Mixed | CEA | Cognistat, FAB | 76.95 | 83 |
| Mlekusch  2008 | Prospective | Mixed | CAS | TMT, Oral word association, Supermarket items, Animal test | 71 | 59.2 |
| Ning 2022 | non-RCT comparative | Asymptomatic | CEA,  CAS | MoCA | 45 | 56 |
| Ning 2024 | Prospective | Mixed | CEA,  CAS | MoCA | 89 | 82 |
| Ogasawara  2005 | Prospective | Mixed | CEA | WAIS, WMS, ROCF | 92 | 88 |
| Oliveira  2014 | non-RCT comparative | Mixed | CEA,  CAS | MMSE. RAVLT, FDS, BDS, Stroop, TMT, VF,  FAS, BNT, LNI | 30 | 76.7 |
| Ortega  2013 | Prospective | Mixed | CAS | WAIS-III, MS-III, BNT, Token test, Controlled  Orgal Word Association, Semantic Fluency,  California Verbal Learning, Grooved  Pegboard, BJLO, Stroop, COWA, | 46 | 84.8 |
| Pearson  2003 | Prospective | Mixed | CEA | TMT, BRVT, RAVLT, | 39 | 73 |
| Pichetto  2013 | Prospective | Asymptomatic | CEA,  CAS | Rey auditory verbal, Phonological verbal fluency, Wisconsin card sorting, Copying drawings | 22 | 63.6 |
| Piegza 2022 | Prospective | Mixed | CAS | Digit Symbol, Rey-Osterrieth complex figure test, Wisconsin Card Sorting test, Letter verbal fluency | 47 | 66 |
| Plessers  2015 | Prospective | Mixed | CEA,  CAS | TMT, Spatial Span, Digit span, Stroop, Symbol substitution, AVLT, Phonological verbal fluency, Semantic verbal fluency, CFT, D-2,  Grooved Pegboard, Line bisection | 72 | 62.5 |
| Plessers  2016 | non-RCT comparative | Asymptomatic | CEA,  CAS | MMSE, | 72 | 64.3 |
| Porcu 2022 | Prospective | Asymptomatic | CEA | MMSE | 20 | 70 |
| Porcu 2022 | Prospective | Asymptomatic | CEA | MMSE | 19 | 68 |
| Pucite 2019 | non-RCT comparative | Mixed | CEA,  CAS | MoCA | 213 | N/A |
| Raabe 2010 | Prospective | Mixed | CAS | DRS-2. BDI-II, NAART, 3MS, MMS, TMT-B,  RAVLT, LES Mayo, DRS Mayo, PRS Mayo | 62 | 67.7 |
| Relander  2024 | non-RCT comparative | Mixed | CEA | Digit Span forwards/backwards WMS-III,  Spatial Span forwards/backwards WMS-III, Cogstate One Back/Two Back test with playing cards, Stroop test, Modified Rey, Stroop color, number sequencing of FAT, Finger tapping,  Purdue Pegboards | 98 | 77 |
| Scherr 2016 | Prospective | Asymptomatic | CAS | MMSE, Semantic verbal fluency, Phonemic verbal fluency, BNT, Word list, Discriminability, Figure Recall, TMT | 72 | 66.7 |
| Shi 2016 | non-RCT comparative study | N/A | CEA,  CAS | MMSE, MoCA, P300 | 235 | 87.2 |
| Skoloudik  2016 | RCT | Mixed | CEA,  CAS | MMSE, Clock-drawing, Verbal fluency | 242 | 68.9 |
| Song 2019 | non-RCT comparative | Symptomatic | CAS | MoCA, Line connection, Copy cube, Drawing clock, Naming, Attention, Sentence repeating, Verbal fluency, Abstraction, Delayed recall, Orientation | 79 | 67.1 |
| Soni 2021 | Prospective | Mixed | CAS | ACE-R | 25 | 88 |
| Takaiwa  2009 | non-RCT comparative | Mixed | CEA,  CAS | MMSE, RBANS | 26 | 76.9 |
| Takaiwa  2013 | Prospective | Asymptomatic | CEA | WAIS-R, JART, RBANS | 15 | 80 |
| Tani 2019 | Prospective | Asymptomatic | CAS | WAIS-III, WMS-R | 8 | 100 |
| Thirumala  2020 | Retrospective | Mixed | CEA | DSST, 3MSE | 167 | 58.7 |
| Turk 2008 | Prospective | Symptomatic | CAS | MMSE, RBANS, TMT, IQCODE | 17 | 88.2 |
| Turowicz  2021 | non-RCT comparative | Asymptomatic | CAS | MoCA, CANTAB | 105 | 63.8 |
| Usman  2016 | Prospective | Mixed | CEA | ACE, GPCOG | 79 | 87.3 |
| Wang 2017 | Prospective | Asymptomatic | CAS | MMSE, MoCA, Verbal memory, Digit span,  Rey auditory verbal learning, Digit symbol | 16 | 75 |
| Wapp 2015 | Prospective | Mixed | CEA,  CAS | Stroop, Symbols, Animal Naming, BNT, Word  Rey Learning, Digit Span, Signs, Rey Figure,  Purdue Pegboard, Hospital Anxiety and  Depression Scale, | 58 | 74.1 |
| Wasser  2011 | non-RCT comparative | Mixed | CEA,  CAS | TAP, WMS, SRT, RWT, WCST, Rey complex rigure, NVLT, SPAT, LGT-3 | 55 | 78.2 |
| Watanabe  2014 | Prospective | Asymptomatic | CEA | MMSE. MoCA | 39 | 91.7 |
| Witt 2007 | non-RCT comparative | Symptomatic | CEA,  CAS | RAVLT, TMT, Rey Complex Figure, PVSAT,  Stroop. Verbal Fluency, RNG, PP-Test, Beck  Depression Inventory, Finger tapping, NIHSS | 45 | 62.2 |
| Wu 2024 | Prospective | Mixed | CAS | MMSE, DTI-ALPS index | 102 | 84 |
| Xu 2007 | non-RCT comparative | Mixed | CAS | MMSE, RAVLT, Rey complex figure, Digit span, TMT, BNT, Finger tapping | 110 | 77.5 |
| Yan 2014 | non-RCT comparative | Mixed | CAS | MoCA, Line connection, Copy cube, Drawing clock, Naming, Attention, Sentence repeating, Verbal fluency, Abstraction, Delayed recall,  Orientation, HAMD, HAMA, WHOQOL-  BREF | 65 | 52.3 |
| Ye 2022 | non-RCT comparative* | Mixed | CEA,  CAS | RVR, Digit Span Test | 90 | 57 |
| Yoon 2015 | non-RCT comparative | 12  Asymptomatic,  11 Symptomatic | CAS | K-MMSE, Digit span, K-BNT, calculation, ROCFT, Orientation, SVLT, ROCFT, Contrasting program, Go-no-go, Fist-edgepalm, Luria loop, Word fluency, Stroop,  SNSB-D | 33 | 75.8 |
| Zhang 2016 | non-RCT comparative | — | CEA | MMSE, Clock drawing | 67 | 70.1 |
| Zhou 2017 | non-RCT comparative | Mixed | CEA,  CAS | RAVLT | 119 | N/A |

**SUPPLEMENTAL FIGURE 1.** Funnel plots for small-study effects analyses. Visual asymmetry should be interpreted cautiously as an indicator of small-study effects rather than definitive publication bias, particularly in strata with limited numbers of studies.

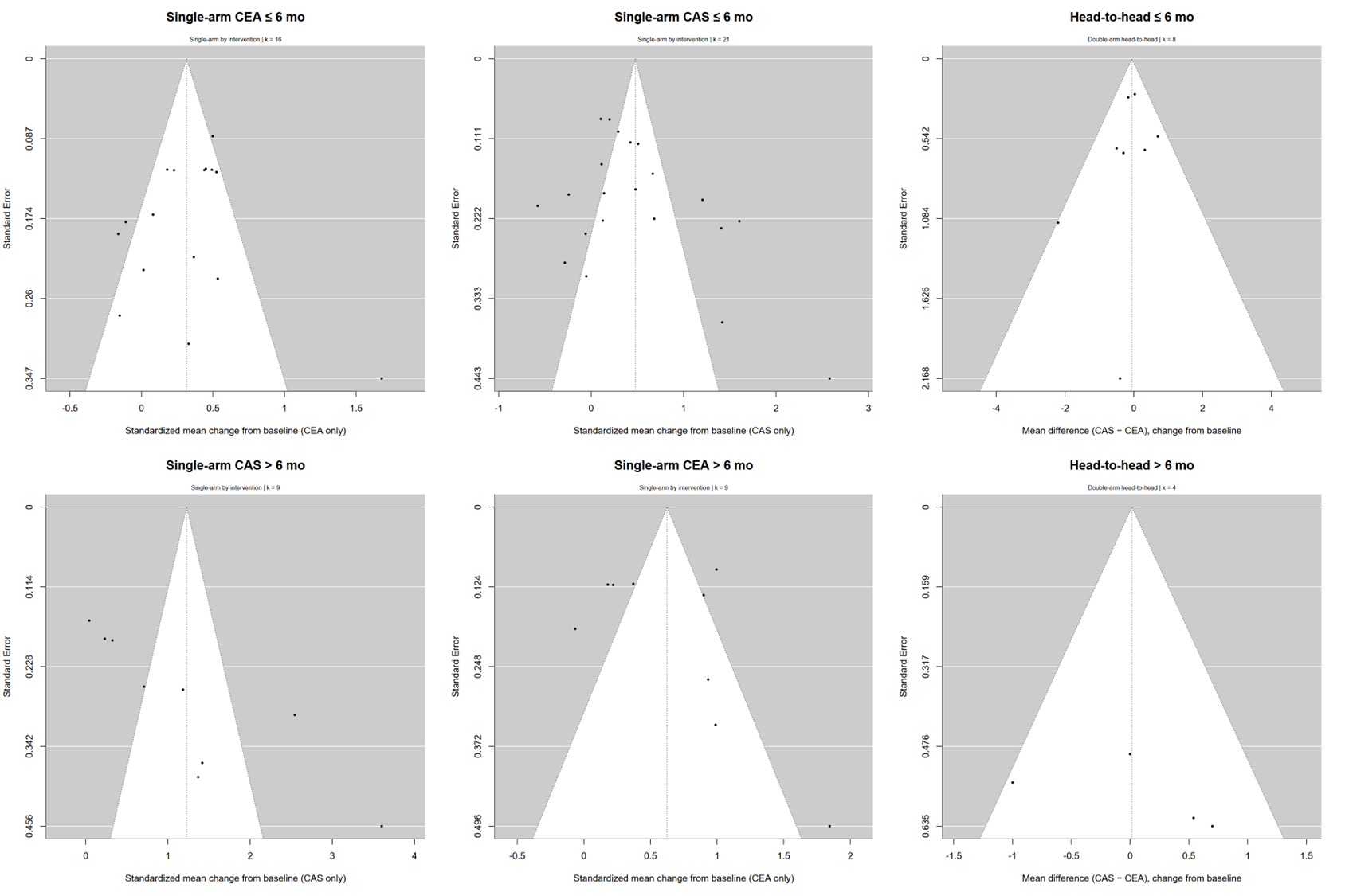

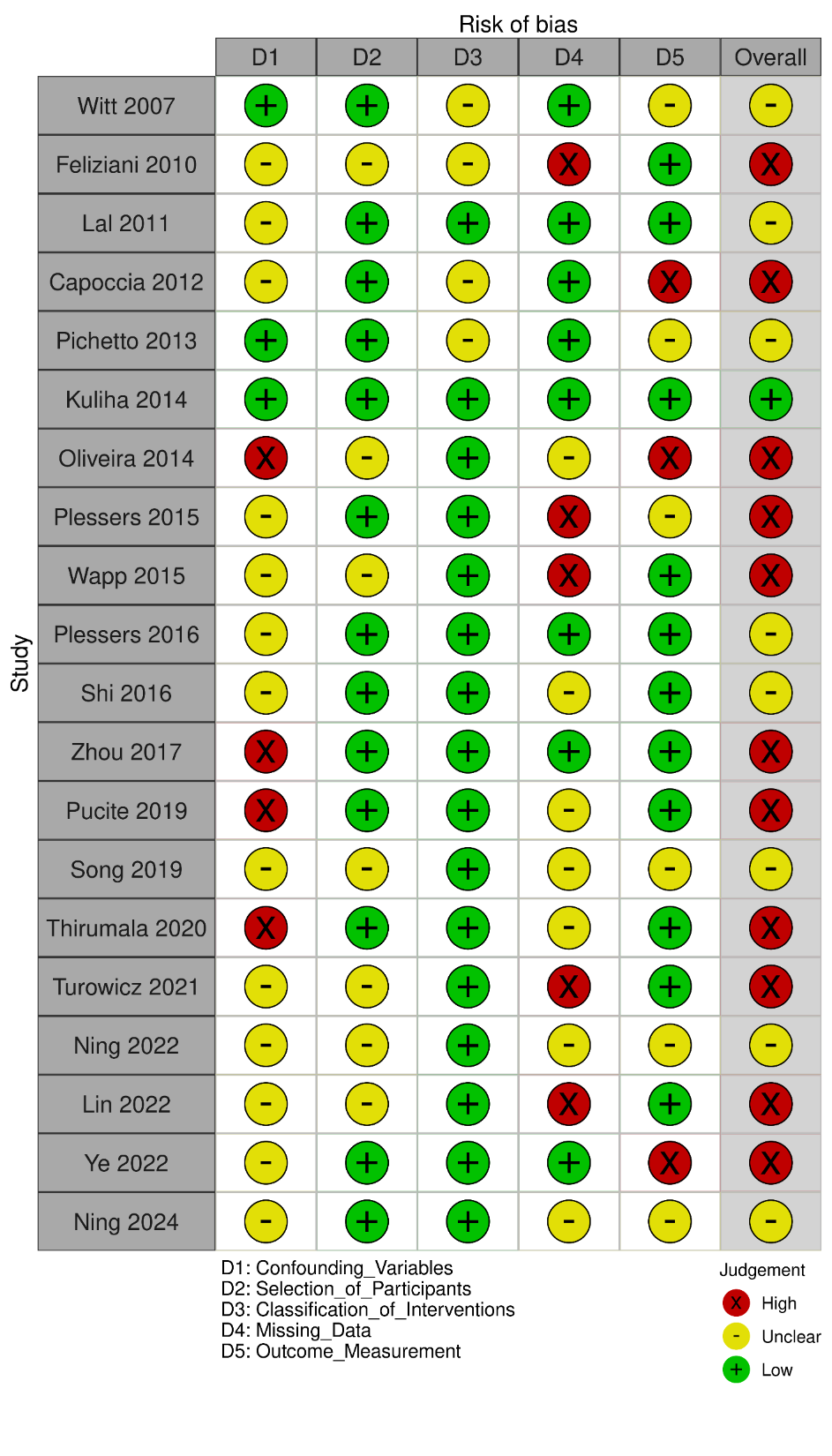

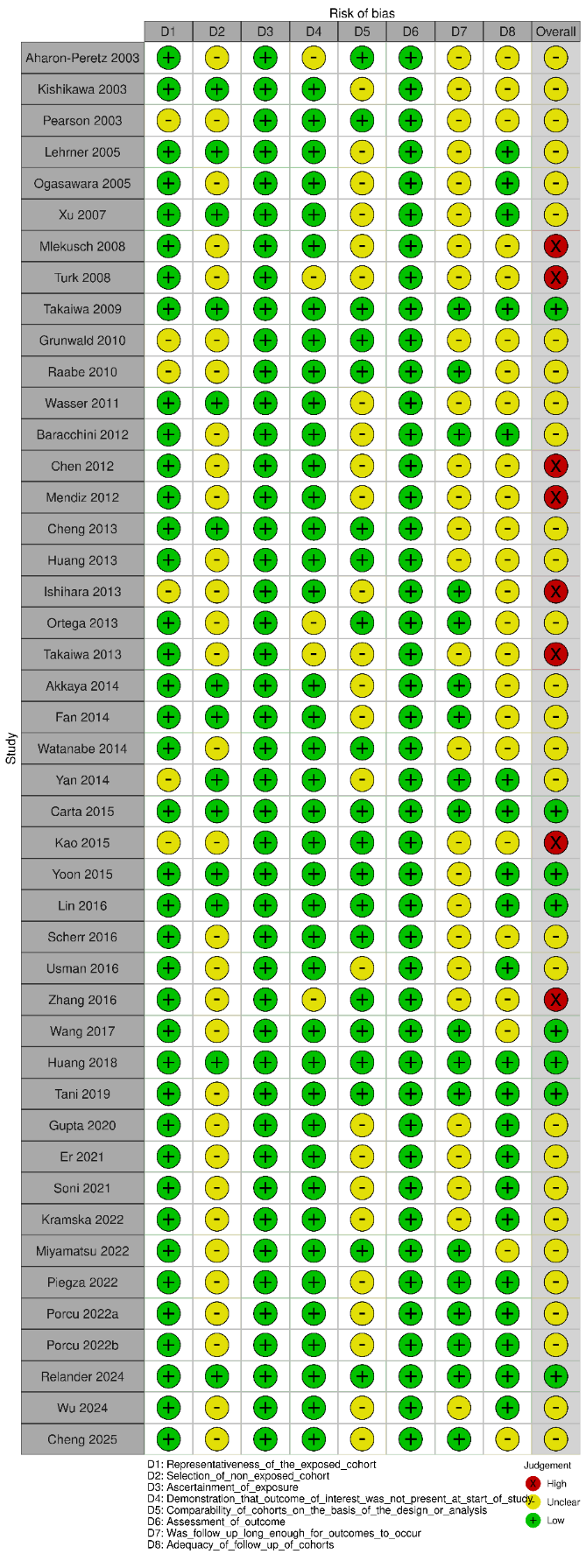
**SUPPLEMENTAL FIGURE 3.** Traffic-light plots of risk of bias analysis. NOS is represented most (left, k = 45), followed by ROBINS-I (right, k = 20) and ROB-2 (below, k = 3).

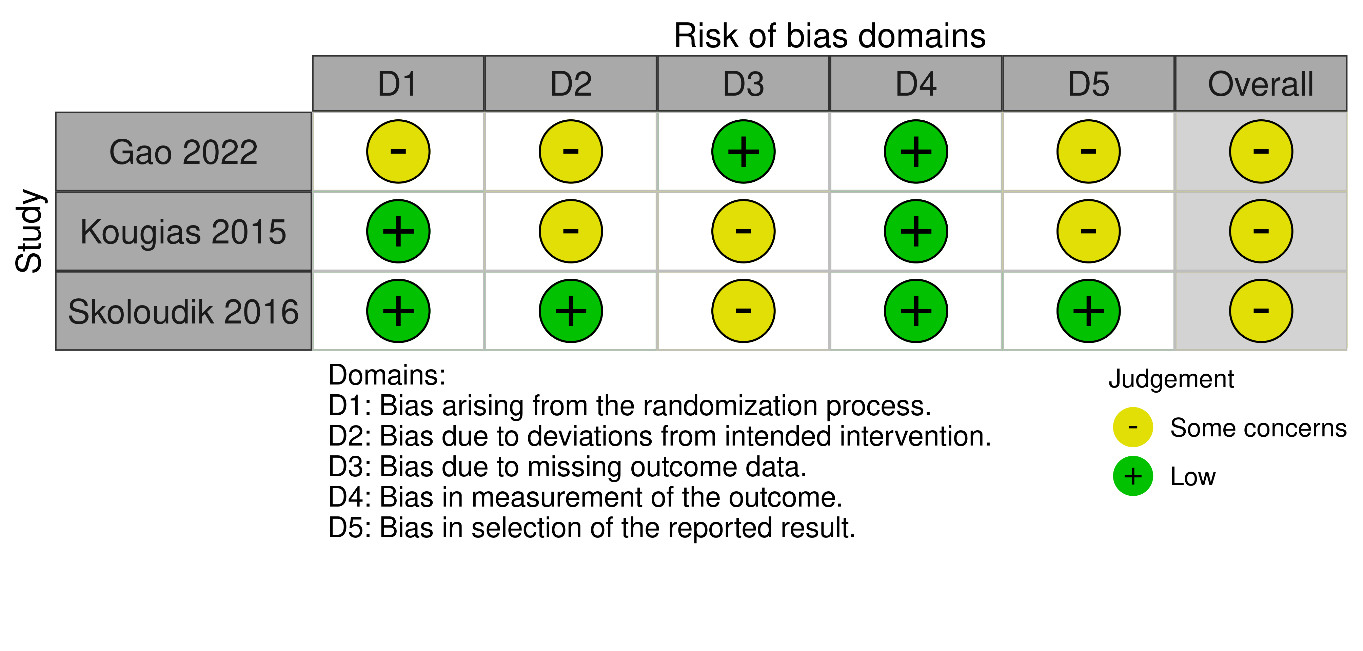
